# Development and external validation of an ultrasound image-based deep learning model to estimate gestational age in the second and third trimesters of pregnancy using data from Garbh-Ini cohort: a prospective cohort study in North Indian population

**DOI:** 10.1101/2024.05.13.24305466

**Authors:** Divyanshu Mishra, Varun Chandramohan, Nikhil Sharma, Mudita Gosain, Nitya Wadhwa, Uma Chandra Mouli Natchu, GARBH-Ini study group, Ashok Khurana, J. Alison Noble, Ramachandran Thiruvengadam, Bapu Koundinya Desiraju, Shinjini Bhatnagar

## Abstract

Accurate estimation of gestational age (GA) is essential to plan appropriate antenatal care. Current GA estimation models rely on fetal biometry measurements, which are susceptible to ethnic and pathological variations in fetal growth, especially in the second and third trimesters of pregnancy. In this study, we challenge the current paradigm of estimating GA using fetal biometry, by using ultrasound (US) images and deep learning models which can automatically learn image features associated with GA. We developed deep learning models for GA estimation using US images taken at 18-32 weeks of pregnancy from 2207 participants of Garbh-Ini - a hospital-based prospective cohort of pregnant women in North India. Further, we designed a novel conformal prediction (CP) algorithm to detect and reject images when there is a data distribution shift, preventing erroneous predictions. Our best model, GArbh-Ini Ultrasound image-based Gestational age Estimator (GAUGE), which was trained on US images of the fetal head (9647 images from 2207 participants), had a mean absolute error (MAE) of 2.8 days when evaluated on an internal test dataset (N = 204). GAUGE is 44% and 35% more accurate than the widely used Hadlock and INTERGROWTH-21st biometry-based GA models, respectively on the internal test dataset. For an external test dataset (N = 311), collected retrospectively from The Ultrasound Lab, New Delhi, the same model achieved a MAE of 5.9 days. In addition, we show that GAUGE relies on the finer details in the image instead of the fetal biometry and that this leads to a similar performance across small for gestational age (SGA) and appropriate for gestational age (AGA) groups. The ability of GAUGE to consider image features beyond derived biometry suggests that GAUGE offers a better choice for populations with a high prevalence of fetal growth restriction.

## 1 Introduction

Gestational age (GA) estimation is the cornerstone of obstetric care, and its significance is widely known(1, 2). GA not only influences clinical decision-making for monitoring fetal growth and other pregnancy related complications, but also affects population-level estimates of pregnancy outcomes such as preterm birth, fetal growth restriction, and stillbirth(3, 4). Traditionally, GA is estimated using the first day of the last menstrual period (LMP). This method has several assumptions and is imprecise in a large proportion of women with recall bias, irregular menstrual periods, oral contraceptive use, and recent breastfeeding(5, 6). Currently preferred ways of estimating GA in clinical practice use ultrasound-based measurements of fetal biometry such as crown rump length (CRL), head circumference, biparietal diameter, occipitofrontal diameter, femur length, and abdominal circumference. In clinical practice, the recommended and accurate way of estimating GA uses CRL measured before14 weeks of gestation(7, 8). GA estimation in the second and third trimesters (> 14 weeks) relies on fetal biometry parameters other than CRL, which are susceptible to ethnic and pathological variations in fetal growth especially in this period(1, 2). In low middle income countries (LMICs) such as India, where nearly 30% of pregnant women seek antenatal care beyond their first trimester(9) and a high prevalence of small for gestational age fetus (35%), the current biometry-based methods are a mere approximation. In such a scenario, the only way of achieving accurate GA estimation, particularly in the latter part of pregnancy, are to rely on fetal anatomies that are spared in growth restriction conditions(10) or to have non-biometry based information in the ultrasound (US) images to determine GA.

Deep learning approaches do not rely on derived biometry and can automatically learn image features associated with GA. Recent studies using deep learning on US images and video sweeps for GA estimation have shown better results than models based on fetal biometry(11–15). Many of these studies use opensource or retrospective data and lack external validation. Another limitation of existing models, in fact all deep learning models, is that they predict on any given input image even when it is out-of-distribution thereby raising concerns on their trustworthiness.

In this paper, we tried to overcome these limitations by using high quality data, collected prospectively. and managed using standard protocols, in Garbh-Ini cohort from North India and demonstrated the model’s performance by externally validating the model. Further, we introduce a novel conformal prediction framework that identifies out-of-distribution samples and prevents erroneous predictions. In summary, we present GArbh-Ini Ultrasound image-based Gestational age Estimator (GAUGE), a deep learning-based model for GA prediction. We showed GAUGE is more accurate than existing biometry-based methods, depends on the finer details in the image, and performs consistently in both SGA and AGA groups.

## 2 Results

### Study population

Of the 8235 participants recruited into the GARBH–Ini cohort, data from 5594 participants, enrolled at <14 weeks of gestation, was considered for the analysis. A total of 833 participants with abortions (18), still birth (119), multiple pregnancy (44), loss to follow-up (652) were excluded. To ensure that the gold standard GA was accurate, we excluded women where the GA calculated using CRL and that using LMP disagreed by over a week (1541). Finally, US images of the head (n=9647), abdomen (n=9342), and femur (n=8285) from 2207 participants were included in the analysis (Fig. 1). The median age of the participants was 23 years (IQR 21, 26), with 21% underweight (body mass index (BMI) < 18.5 kg/m^2^) and 14.1% overweight (25 kg/m^2^ <= BMI < 30 kg/m^2^) or obese (BMI >= 30 kg/m^2^). More than half (51%) of the participants were nulliparous. The median fetal gestational age at enrollment into the study was 10.1 weeks (IQR 8– 12.1 weeks). The detailed sociodemographic and clinical characteristics of the included participants are summarized in Table 1. The comparison of baseline characteristics show that the participants included in this study were representative of all participants of the parent cohort (Supplementary Table 1).

**Fig 1:**
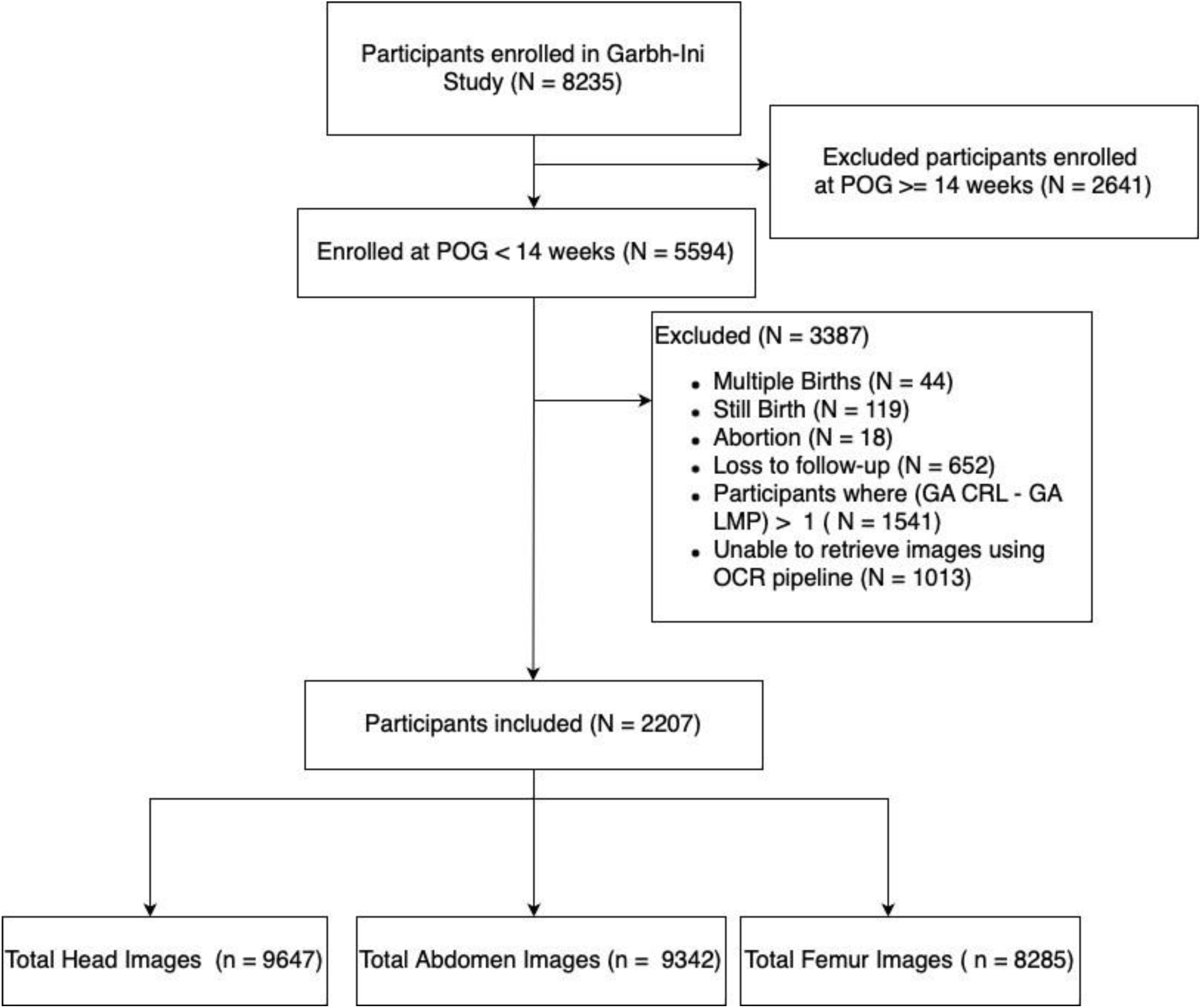
Study Flow. Flowchart showing the number of participants enrolled in GARBH-Ini cohort and images considered for analyses. *POG: Period of gestation

**Table 1:** Clinical and sociodemographic characteristics of participants included in the analysis (N = 2207)

| **Variables** | **Median (IQR) or N (%)** |
| --- | --- |
| Age | 23.0 (21.0, 26.0) |
| Body Mass Index (BMI)<br>Underweight<br>Normal<br>Overweight<br>Obese | 465 (21%)<br>1,429 (65%)<br>255 (12%)<br>45 (2.1%) |
| Gestational age at enrolment | 10.1 (8.0, 12.1) |
| Parity:<br>Nulliparous<br>Parous | 1,122 (51%)<br>1,072 (49%) |
| Appropriate for gestational age (AGA)<br>Small for gestational age (SGA)<br>Large for gestational age (LGA) | 1,173 (62%)<br>665 (35%)<br>45 (2.4%) |
| Preterm Birth<br>Term Birth | 275 (13%)<br>1,820 (87%) |
| Occupation:<br>Unemployed<br>Employed | 2,014 (92%)<br>179 (8%) |
| Education:<br>Illiterate<br>Literate | 406 (19%)<br>1,788 (81%) |

### Fetal head images were sufficient for accurate modelling

Initially, we developed three separate CNN regression models using the US images of the head, femur, and abdomen, respectively to identify which anatomy is best to use in a model. The model trained using images of the head was most accurate with a MAE of 3.5 days, compared to those that used images of the femur (MAE:6.3 days) and abdomen (MAE:7.1 days) on the internal test dataset (Supplementary Table 2). We have also developed a multi-input model which takes all three images (head, femur, and abdomen) and predicts GA to evaluate if this combination of images can improve the prediction. Contrary to our expectations, there was no improvement in the prediction (MAE of 3.6 days). These results suggest that images of the fetal head harbor all the information required to predict GA and are sufficient to build accurate models. Based on these observations, we used only fetal head images for further modelling.

### Development and validation of GAUGE model

In addition to the baseline regression model based on head images, we designed a novel multi-task model (GAUGE) that segments the head along with GA estimation (Fig. 2a). The MAE varied by less than a day during bootstrapping (30 runs) demonstrating the good convergence of our model. The samples which were rejected by our CP framework (Fig. 2b) have higher error rates in both internal and external validation datasets substantiating its use while predicting on new samples (Supplementary Fig. 1). The CP framework identified images from 7.7% participants from the internal test and 62% from the external test set as out-of-distribution samples and rejected them before the prediction (Supplementary Fig. 2, Supplementary Table 3). The GAUGE model had an overall MAE of 2.8 days on the internal test dataset and was chosen as the final model for GA estimation. In the 18-20 (329 images), 20-30 (30 images) and 30+ week (475 images) windows, the MAE of the GAUGE model was 2.3, 5.6, and 3.0 days respectively. When compared with existing biometry-based models, the GAUGE model was 44% (MAE in days 5.11 vs 2.8) and 35% (MAE in days 4.41 vs 2.8) more accurate than the widely used Hadlock(16) and Intergrowth-21st(2) GA models respectively on our internal test dataset (Fig. 3). On the external test dataset, the GAUGE model achieved an overall MAE of 5.9 days and had an MAE of 4.1, 8.4, and 4.9 days in 18-20, 20-30 and 30+ week windows respectively (Table 2).

**Fig 2:**
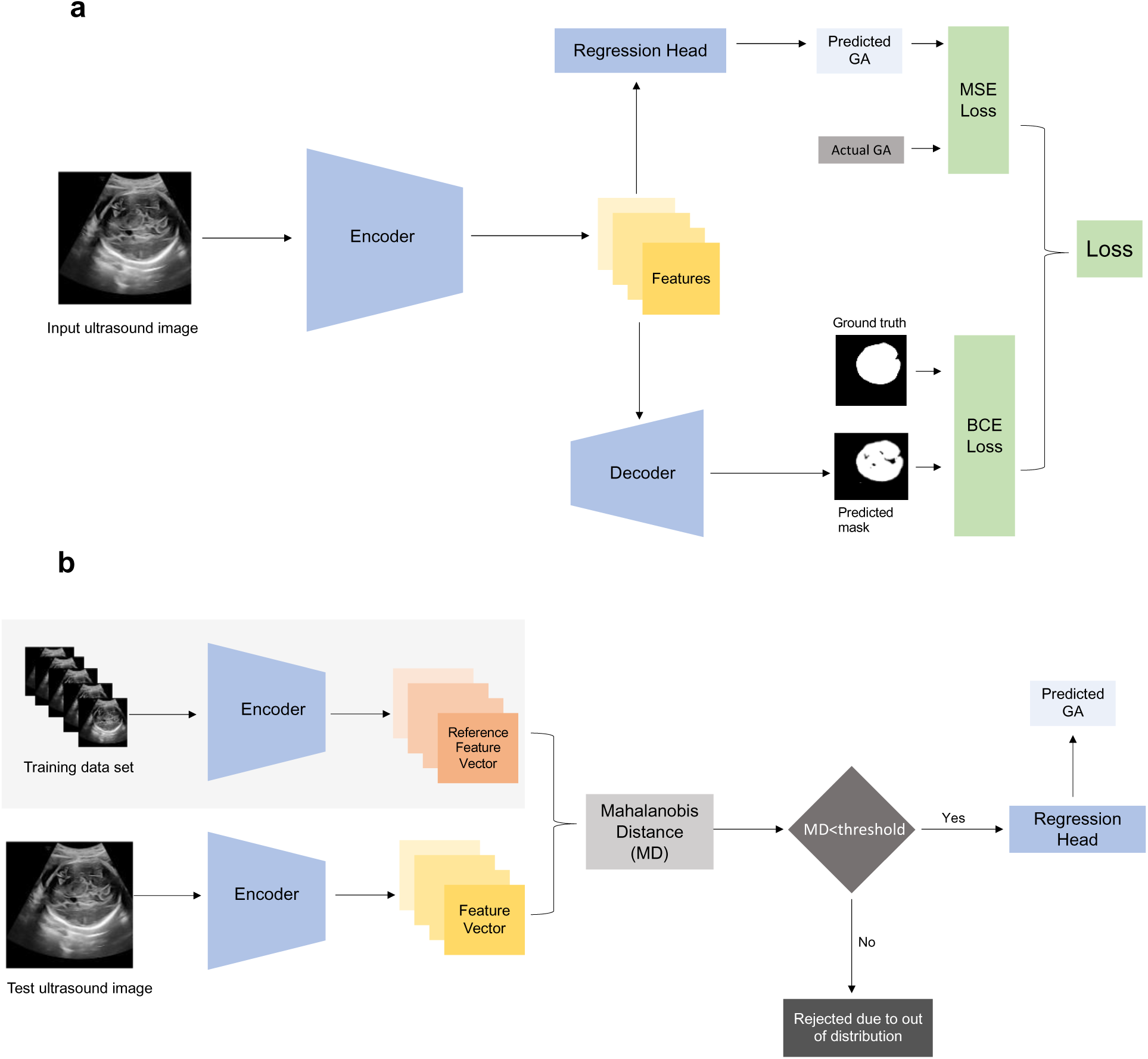
Schematic diagram of GAUGE. **a** shows architecture of GAUGE. The encoder extract features from the input images which are used by decoder to segment the head region and by regression head for GA prediction. The losses of both tasks were combined to calculate a total loss which was used to optimize the model. **b** Working of the conformal prediction framework during inference. The reference feature vector was calculated by averaging the feature vectors of all the samples in the training dataset (part of the image shown in grey background). During inference, the feature vector for each image is compared to the reference feature vector and Mahalanobis Distance (MD) between them is calculated. Lastly accept/reject decision is made by comparing MD of the new image to distribution of MD’s calculated on training data.

**Fig. 3:**
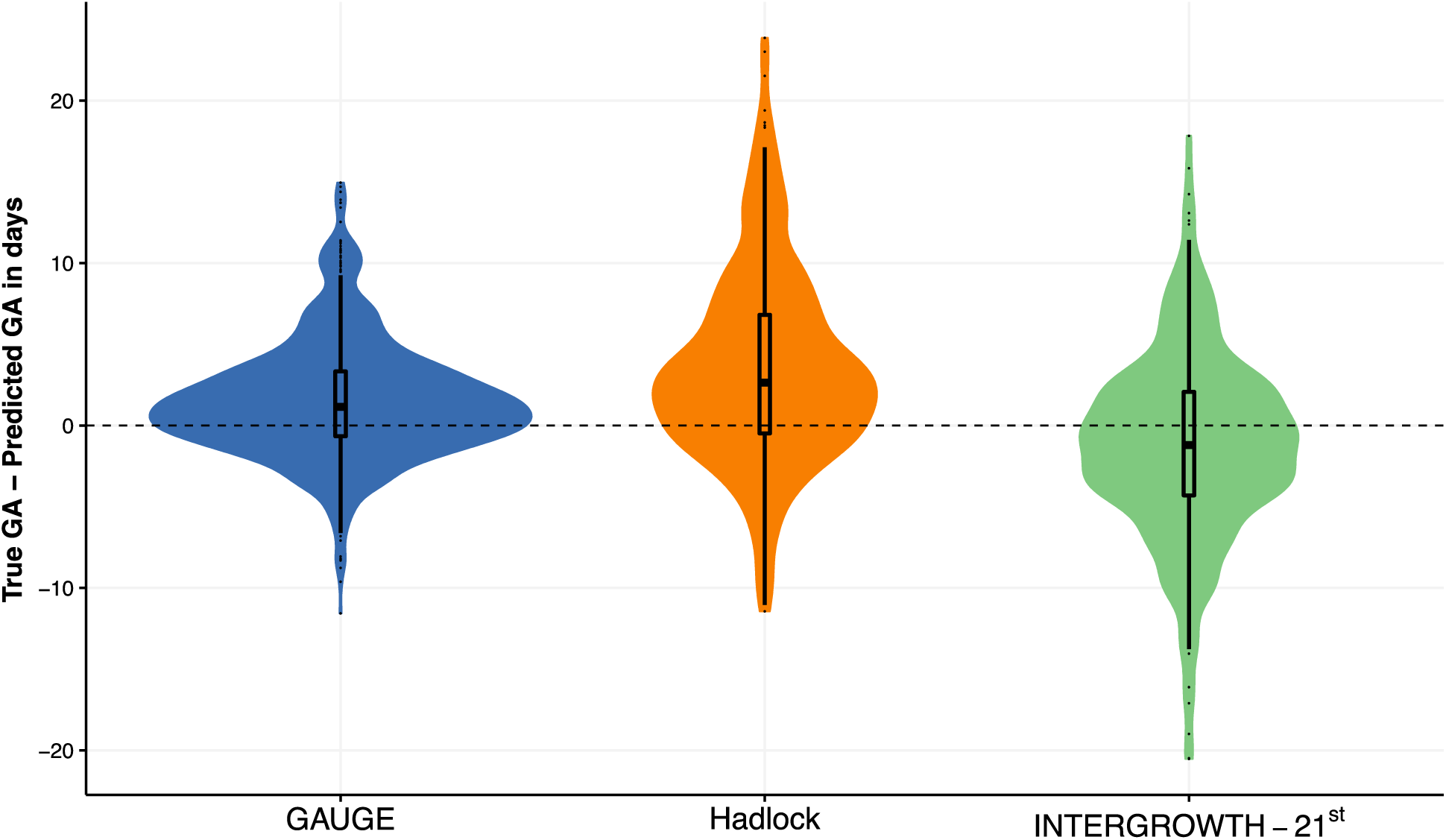
Comparison of error distributions of GAUGE, Hadlock, and INTERGROWTH-21^st^ models (N = 204). Violin and box plots showing the distribution of error in days. The error of GAUGE is centered around zero and a smaller range when compared to the biometry-based methods of Hadlock and INTERGROWTH-21^st^.

**Table 2:**
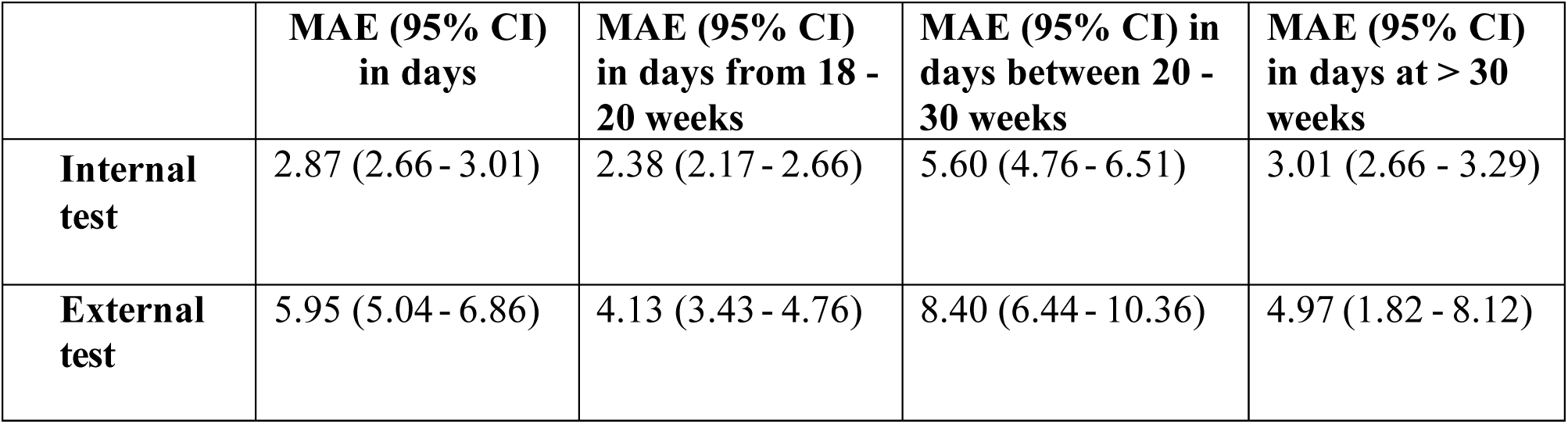
Accuracy of the GAUGE model on the internal and external test datasets reported as mean absolute error and 95% confidence interval in days.

### GAUGE model relies on the finer details in the image

Heat maps were created using a regression activation map technique (Supplementary Fig. 3). These showed that the finer details within the skull were more predictive of GA when compared to the outline of the skull (from which fetal biometry is calculated). To further investigate this finding, we used image blurring and resizing to compromise the finer details in the image while maintaining the outline of the skull. When tested on the images that were blurred using Gaussian filters of sizes 3x3, 5x5, 7x7 and 9x9 pixels, the MAE of the GAUGE model increased to 5.62, 7.96, 9.16, and 9.48 days respectively (Fig. 4, Supplementary Fig. 3). A similar increasing trend in MAE was found (4.81, 7.21 and 12.26 days) when the model was tested on images where the spatial resolution was reduced by 75%, 50% and 25% respectively (Fig. 4, Supplementary Fig. 4). The models which were trained end-to-end on the resized images also showed increased MAE when compared to the GAUGE model (5.9 days). The above results show that fine intensity patterns play an important role in model prediction. This observation is particularly relevant to GA prediction in fetal growth restriction, where existing biometric-based models tend to have high error rates. To investigate this, the errors of the GAUGE, Hadlock and INTERGROWTH-21^st^ models were compared in small for gestational age (SGA) and appropriate for gestational age (AGA) groups. We found that the GAUGE model performed consistently in SGA and AGA groups whereas both the Hadlock and INTERGROWTH-21st models underestimated the gestational age in the small for gestational age as compared to appropriate for gestation babies (Fig. 5, Supplementary Table 4).

**Fig. 4:**
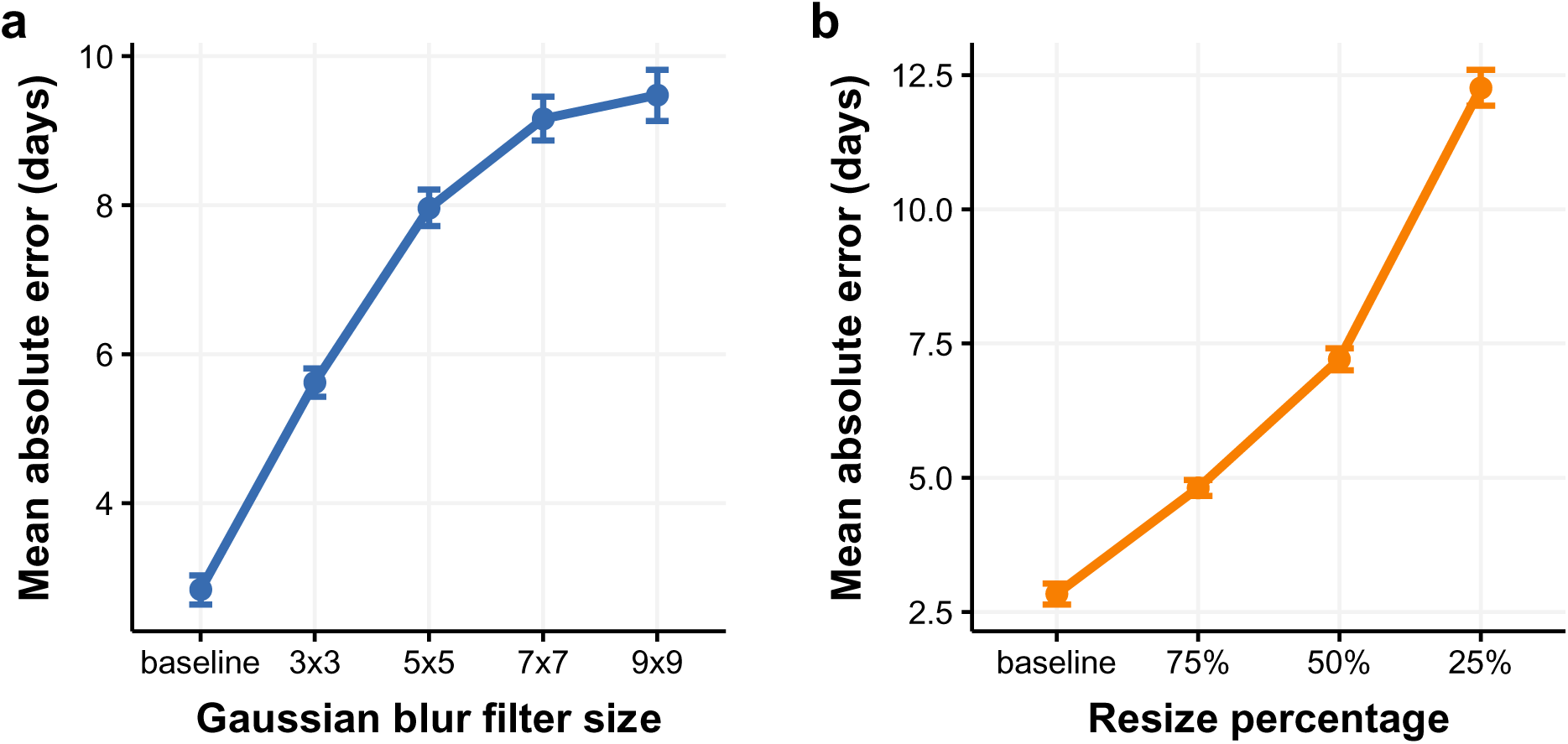
Experiments demonstrating that the model is dependent on the finer details in the image. **a** shows the mean absolute error (MAE) in days and 95% confidence interval when the input images are increasingly blurred using a Gaussian filter of different sizes compared to the baseline (unaltered input image). **b** shows the mean absolute error (MAE) in days and 95% confidence interval when the input images are reduced in size in different proportions compared to baseline (unaltered input image).

**Fig. 5:**
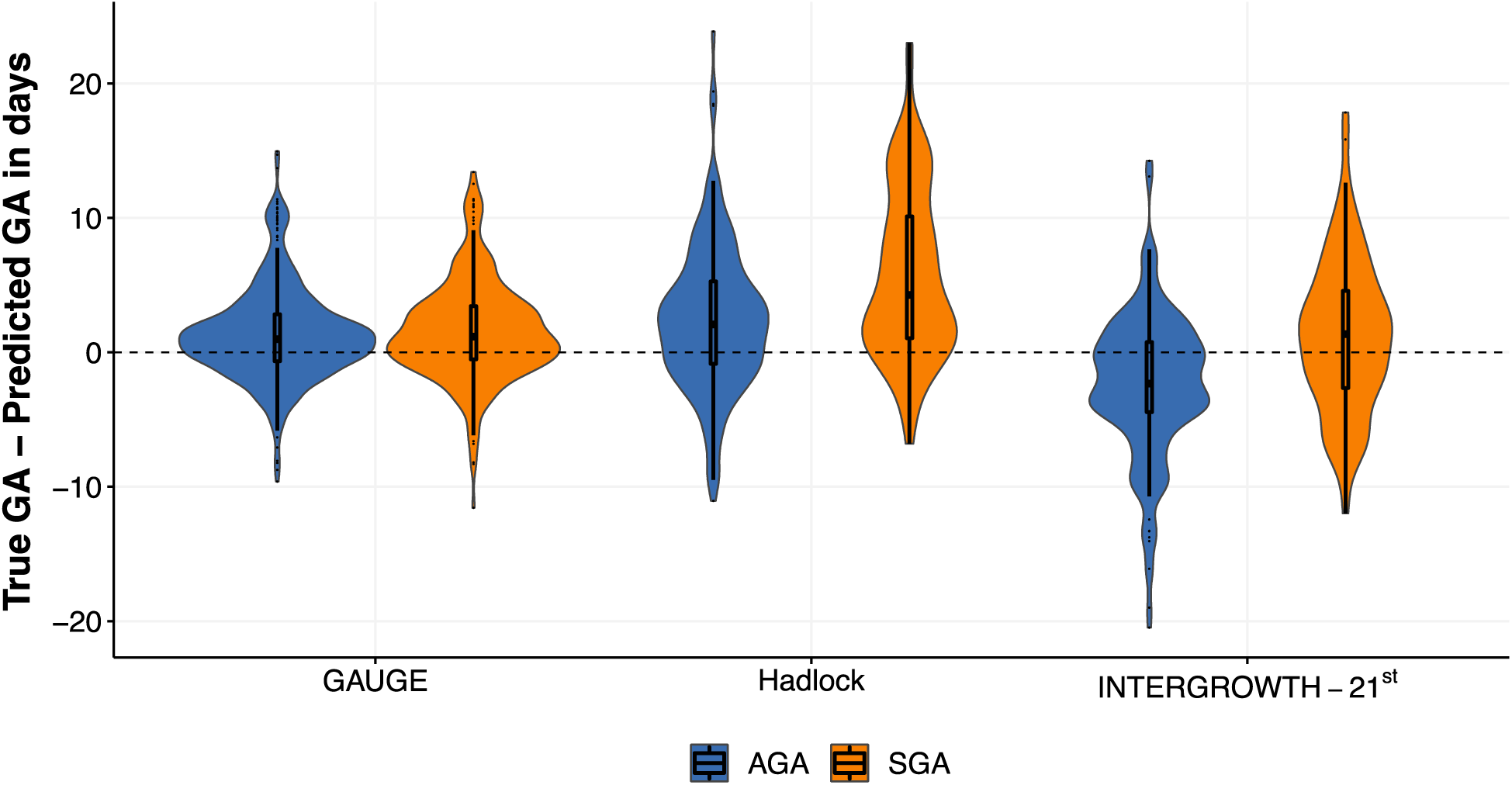
Comparison of error distributions between small for gestational age (SGA) and appropriate for gestational age (AGA) groups (N = 204). GAUGE shows similar performance for both SGA and AGA groups (p = 0.1), while both Hadlock (p < 0.001) and INTERGROWTH-21^st^ (p <0.001) models have underestimated GA in the SGA group when compared to the AGA group. A Kolmogorov Smirnov test was used to compare the distributions.

## 3 Discussion

In this study we developed and externally validated the GAUGE model which predicts GA from the US images of the fetal head in the second and third trimesters of pregnancy. The results of GAUGE model (2.8 and 5.9 days in internal and external validation) are comparable to the two recently published models by Dan *et al.* (MAE of 5 days) and Lee *et al.* (MAE 3 days in second trimester and 4.3 days in third trimester). While Dan *et al*. used a large hospital based retrospective data, Lee *et al*. used data from the prospective cohort and performed external validation like our study. We challenged the current clinical paradigm of estimating GA using fetal biometry and demonstrated that GAUGE performs consistently in SGA and AGA populations unlike biometry-based models. Existing clinical GA estimation models exclusively use fetal growth measurements and are error-prone in populations with a high prevalence of abnormal fetal growth patterns(2, 10, 12). Our model convincingly addresses the limitations of existing clinical GA estimation models and is appropriate for use in such populations.

The architecture of the GAUGE model is designed to focus on the features relevant to the fetal head for GA estimation (Fig. 2a). Although segmentation of the fetal head is separate from the prediction of gestational age and is not essential, we found that this strategy enabled us to achieve better accuracy and generalization than the simple image regression model. (Supplementary Table 5). Additionally, our model provides a visual representation of the region of the fetal head which would inspire more confidence on the model amongst users although further research is critical. Despite performing two tasks, GAUGE model has similar computational requirements (26.1 million parameters) when compared to a simple image regression model (21.3 million parameters).

One of the concerns recognized in using deep learning models is the reliability of prediction on new samples (for example: samples from diverse geographical regions, acquired using different machines, and with varied resolutions and quality). Inspired by recent literature(17), we have introduced a novel image-based conformal prediction framework to prevent GAUGE from predicting on data where there is a distribution shift relative to the data used to train the model, thereby preventing erroneous predictions. Our intention here is to detect the out-of-distribution data and reject it. Our approach is different from the domain adaptation techniques, where the goal is to adapt the model by using the out-of-distribution data. We hypothesize that the ability of our model to say “I don’t know” increases trust in the model predictions and the same can be evaluated in future studies.

One of the limitations of our work is that there were fewer samples in our training and test datasets between 20–30 weeks of gestation compared to 18-20 or 30 + weeks owing to the ultrasound scan schedule for the GARBH-Ini cohort which aligned with the antenatal standard of care available to the participants. This is also reflected in the wider confidence interval of MAE during this window. Further, we found that half of the samples from the external test dataset were rejected by the conformal prediction step. We believe that this is due to the lower spatial resolution images in the external test dataset when compared to training images (240 x 320 pixels vs 1920 x1080 pixels). Other reasons for rejection by the conformal prediction algorithm could be because of participant characteristics, and different US machines.

We have developed deep learning models for GA estimation that are more accurate than those used in current routine clinical practice. This may be due to our state-of-the-art deep learning models and training techniques, a large dataset from a prospective cohort, and an accurate gold standard. As we are using the images directly as input to the machine learning model rather than using the measured values of the fetal biometry, an additional potential advantage is that GAUGE reduces time, effort, and the inter-operator variability which may occur during the manual measurement of fetal biometry in US images. From the perspective of translating this model to clinical practice, our next steps are to evaluate the GAUGE model in diverse settings for its accuracy, to characterize the image rejection and to package the model within a software tool that seamlessly integrates into the sonologists’ workflow. Overall, we believe that our GAUGE model has the potential to improve antenatal care and subsequently perinatal health outcomes particularly in LIMC settings.

## 4 Methods

### GARBH–Ini Cohort description

This study is nested within the ongoing GARBH–Ini (interdisciplinary Group for Advanced Research on BirtH outcomes-DBT India Initiative) cohort at Gurugram Civil Hospital (GCH), Haryana, India, initiated in May 2015. Pregnant women are enrolled in this ongoing prospective observational cohort before 20 weeks of gestation based on a dating US scan. . The participants are scheduled for four follow-up antenatal visits for an US scan during weeks 11–14, 18–20, 30–32 and 35-37. Socio-demographic, environmental and clinical information is collected at the time of enrollment and during follow-up visits by trained medical professionals using questionnaires that follow standard protocols(18).

Images are acquired by a study radiologist using a GE Voluson E8 Expert (General Electric Healthcare, Chicago, Illinois) and GE Voluson E8 ultrasound machine. For each participant, the radiologist captures images required for the fetal biometry in triplicates according to standard protocols (harmonized with Alliance for Maternal and Newborn Health Improvement protocols(19)). Twenty percent of randomly selected de-identified images are reviewed by a team of experienced radiologists every month for quality monitoring using a scoring system (18). The detailed methodology on the image acquisition, management, quality assessment protocols are published elsewhere. All the images are de-identified and stored in a server located at the Translational Health Science and Technology Institute, Faridabad. Further details of the methodology are published previously(18).

### Ethics and Informed Consent

Our study was approved by the ethics committee of the Gurugram civil hospital, Gurugram and Translational Health Science and Technology Institute, Faridabad and adhered to the tenets of the Declaration of Helsinki as part of the overall Garbhini cohort objectives. Written informed consent was obtained from the participants after the nature of the study was explained. If an eligible woman was illiterate, a thumb impression was taken from her after ensuring that she had understood and had also explicitly stated her consent verbally; a literate impartial witness signed the consent form.

### Image acquisition and dataset preparation

The US images of the head, abdomen and femur collected during the second and third trimesters were extracted from the image repository using OCR-based software developed in-house (detailed in supplementary methods). The images segregated by the OCR software had text annotations on them which are not suitable for building machine learning-based models as they could introduce model bias. We cropped the images to remove the biometry values on the bottom right of the images using a deep learning-based cropping model. Annotations on the region of interest were removed by image inpainting. These preprocessing steps are explained in detail in supplementary methods section.

### Ground truth calculation

The GA derived from Hadlock formula using CRL at < 14 weeks of pregnancy was considered as the GA gold standard and was available for all training and test images. For image segmentation task, the images of fetal head were manually annotated using Computer Vision Annotation Tool to generate the ground truth.

### GA estimation models

Estimating GA from a US image is treated as an image regression task and to model it, we modified the ResNet 34 architecture(20) by replacing the classification layer with a regression layer. We trained three separate models with the above-mentioned architecture on second and third trimester US images of the head, abdomen, and femur images.

### GAUGE

For developing the GAUGE model we selected only those GARBH-Ini cohort participants who were enrolled at < 14 weeks and for whom CRL based dating using Hadlock formula was available. GAUGE has a UNET++ segmentation architecture(21) at its core. To predict GA using the same architecture we have modified it by adding a separate regression head which utilizes the features of the UNET++ encoder. The encoder and decoder of the model have a ResNet34 backbone and use an AdamW optimizer to optimize a combination of Binary Cross Entropy and Mean Squared Error Loss for the task of image segmentation and regression respectively.

### Model training procedure

Ten percent of the total dataset was kept aside for final internal testing of the models. The remaining data was split randomly 30 times (bootstrapping) into training (90%) and test (10%) sets. The dataset has three images per participant. We used all three images while training and internal testing. While randomly splitting we used participant id and ensured that all the images of the same participant were present in the either training set or the test set. The external validation dataset has only one image per participant. The images were normalized using the mean and standard deviation of the entire dataset and histogram equalization, and contrast enhancement augmentations were randomly applied. Each model was trained for 100 epochs with a batch size of 128 and model MAE was calculated on the test set. The mean and standard deviation of RMSE on the test set were reported. After all the 30 runs were complete, the model having the best RMSE on the test set was evaluated on the held-out internal and external test sets.

### Computational requirements

All the models were trained in Python 3.8.12 using the Fastai 2.3.0, Pytorch Lighting, and Pytorch 3.9.12 deep learning libraries. The models were trained on a Linux based server having a 72 core Intel Xeon processor, 256 GB of RAM and used a single Nvidia RTX 2080 Ti GPU with 11GB VRAM.

### Conformal prediction

For conformal prediction, first, feature vectors for all the training examples were extracted by passing them through the encoder. The vectors obtained were averaged to calculate a reference feature vector. The Mahalanobis distance was calculated between the reference feature vector and the feature vector of every sample in the training dataset. All these distances were transformed using 1/X^2^ to achieve a normal distribution and were referred as transformed Mahalanobis distance (Supplementary Fig. 5). During inference, any sample that had a transformed Mahalanobis distance (calculated between the reference feature vector and the sample feature vector) less than mean - 1.96(SD) was labelled an out of distribution sample and was not used for prediction.

### Statistical analysis

All continuous variables were summarized using median and interquartile range and categorical variables were summarized using percentage. Mean absolute error (MAE) and root mean squared error (RMSE) were calculated using model predictions and actual values. Bootstrapping (100 runs) was used to calculate confidence interval of MAE. Model errors were visualized by violin plots and Kolmogorov–Smirnov test was used to compare error distributions between the groups. All statistical analyses were performed using R programming language version 4.0.4.

## Data Availability

Data described in the manuscript will be made available upon request pending approval.

## Acknowledgements

**MEMBERS OF GARBH–Ini (in alphabetical order:** Translational Health Science and Technology Institute, NCR Biotech Cluster, Faridabad, India-Coordinating Institute (Shinjini Bhatnagar (PI), Vineeta Bal, Bhabatosh Das, Mahadev Dash, Bapu Koundinya Desiraju, Pallavi Kshetrapal, Sumit Misra, Uma Chandra Mouli Natchu, Satyajit Rath, Kanika Sachdeva, Dharmendra Sharma, Amanpreet Singh, Shailaja Sopory, Ramachandran Thiruvengadam, Nitya Wadhwa); National Institute of Biomedical Genomics, Kalyani, West Bengal, India (Arindam Maitra, Partha P Majumder (Co-PI) Souvik Mukherjee); Regional Centre for Biotechnology, NCR Biotech Cluster, Faridabad, India (Tushar K Maiti); Clinical Development Services Agency, Translational Health Science and Technology Institute, NCR-Biotech Cluster, Faridabad, India (Monika Bahl, Shubra Bansal); Guru-gram Civil Hospital, Haryana, India (Umesh Mehta, Sunita Sharma, Brahmdeep Sindhu); Safdarjung Hospital, New Delhi, India (Sugandha Arya, Rekha Bharti, Harish Chellani, Pratima Mittal); Maulana Azad Medical College, New Delhi, India (Anju Garg, Siddharth Ramji), The Ultrasound Lab, Defence Colony, New Delhi, India (Ashok Khurana); Hamdard Institute of Medical Sciences and Research, Jamia Hamdard University, New Delhi, India (Reva Tripathi); All India Institute of Medical Sciences, New Delhi, India (Alpesh Goyal, Yashdeep Gupta, Smriti Hari, Nikhil Tandon); Government of Haryana, India (Rakesh Gupta); International Centre For Genetic Engineering and Biotechnology, New Delhi, India (Dinakar M Salunke Co-PI); G Balakrish Nair (Rajiv Gandhi Centre for Biotechnology, Trivandrum); Gagandeep Kang (Christian Medical College, Vellore).

This work was funded by DBT Wellcome Trust India Alliance (IA/CPHE/18/1/503947) and Department of Biotechnology, Ministry of Science and Technology, Govt. of India (BT/PR9983/MED/97/194/2013). Professor M. K. Bhan will always be remembered reverently for his critical scientific and technical feedback. The authors recognize the efforts of the personnel of the GARBH-Ini Cohort. The authors are thankful to Mr. Sukdev Sinha and Dr. Anamika Gambhir their scientific support.

## Availability of data and material

Data described in the manuscript will be made available upon request pending approval.

## Code availability

Analysis code will be made available upon request pending approval.

## Authors’ contributions

SB, UCM, NW, RT, AK and GARBH-Ini study team: conducted the GARBH–Ini cohort study and acquired the data; BKD, RT and SB conceived and designed the secondary analysis; DM, VC and BKD analysed data and built AI models; NS has performed statistical analysis; MG and MJ have collected and managed the data and performed quality control. AK collected and contributed to external validation dataset. SB, AN, RT, UCM, and NW guided and provided feedback on the analysis and interpretation of results; DM, BKD, RT and SB wrote the paper. All authors critically reviewed and approved the final manuscript.

## References

1. Cooper S, Somerset D. Determination of Gestational Age by Ultrasound. J Obstet Gynaecol Can. 2016;38(4):337.

2. Papageorghiou AT, Kemp B, Stones W, Ohuma EO, Kennedy SH, Purwar M, et al. Ultrasound-based gestational-age estimation in late pregnancy. Ultrasound Obstet Gynecol. 2016;48(6):719–26.

3. Chawanpaiboon S, Vogel JP, Moller AB, Lumbiganon P, Petzold M, Hogan D, et al. Global, regional, and national estimates of levels of preterm birth in 2014: a systematic review and modelling analysis. Lancet Glob Health. 2019;7(1):e37–e46.

4. Newnham JP, Kemp MW, White SW, Arrese CA, Hart RJ, Keelan JA. Applying Precision Public Health to Prevent Preterm Birth. Front Public Health. 2017;5:66.

5. Creinin MD, Keverline S, Meyn LA. How regular is regular? An analysis of menstrual cycle regularity. Contraception. 2004;70(4):289–92.

6. Cole LA, Ladner DG, Byrn FW. The normal variabilities of the menstrual cycle. Fertil Steril. 2009;91(2):522–7.

7. Salomon LJ, Alfirevic Z, Da Silva Costa F, Deter RL, Figueras F, Ghi T, et al. ISUOG Practice Guidelines: ultrasound assessment of fetal biometry and growth. Ultrasound Obstet Gynecol. 2019;53(6):715–23.

8. Vijayram R, Damaraju N, Xavier A, Desiraju BK, Thiruvengadam R, Misra S, et al. Comparison of first trimester dating methods for gestational age estimation and their implication on preterm birth classification in a North Indian cohort. BMC Pregnancy Childbirth. 2021;21(1):343.

9. Ministry of Health & Family Welfare GoI. National Family Health Survey (https://main.mohfw.gov.in/sites/default/files/NFHS-5_Phase-II_0.pdf). 2019-21.

10. Maternal WHOAf, Newborn Health Improvement Late Pregnancy Dating Study G. Performance of late pregnancy biometry for gestational age dating in low-income and middle-income countries: a prospective, multicountry, population-based cohort study from the WHO Alliance for Maternal and Newborn Health Improvement (AMANHI) Study Group. Lancet Glob Health. 2020;8(4):e545–e54.

11. Lee C, Willis A, Chen C, Sieniek M, Watters A, Stetson B, et al. Development of a Machine Learning Model for Sonographic Assessment of Gestational Age. JAMA Netw Open. 2023;6(1):e2248685.

12. Burgos-Artizzu XP, Coronado-Gutierrez D, Valenzuela-Alcaraz B, Vellve K, Eixarch E, Crispi F, et al. Analysis of maturation features in fetal brain ultrasound via artificial intelligence for the estimation of gestational age. Am J Obstet Gynecol MFM. 2021;3(6):100462.

13. Gomes RG, Vwalika B, Lee C, Willis A, Sieniek M, Price JT, et al. A mobile-optimized artificial intelligence system for gestational age and fetal malpresentation assessment. Commun Med (Lond). 2022;2:128.

14. Dan T, Chen X, He M, Guo H, He X, Chen J, et al. DeepGA for automatically estimating fetal gestational age through ultrasound imaging. Artif Intell Med. 2023;135:102453.

15. Lee LH, Bradburn E, Craik R, Yaqub M, Norris SA, Ismail LC, et al. Machine learning for accurate estimation of fetal gestational age based on ultrasound images. NPJ Digit Med. 2023;6(1):36.

16. Hadlock FP, Deter RL, Harrist RB, Park SK. Estimating fetal age: computer-assisted analysis of multiple fetal growth parameters. Radiology. 1984;152(2):497–501.

17. Shashikumar SP, Wardi G, Malhotra A, Nemati S. Artificial intelligence sepsis prediction algorithm learns to say “I don’t know”. NPJ Digit Med. 2021;4(1):134.

18. Bhatnagar S, Majumder PP, Salunke DM, Interdisciplinary Group for Advanced Research on Birth Outcomes DBTII. A Pregnancy Cohort to Study Multidimensional Correlates of Preterm Birth in India: Study Design, Implementation, and Baseline Characteristics of the Participants. Am J Epidemiol. 2019;188(4):621–31.

19. Amanhi, Baqui A, Ahmed P, Dasgupta SK, Begum N, Rahman M, et al. Development and validation of a simplified algorithm for neonatal gestational age assessment - protocol for the Alliance for Maternal Newborn Health Improvement (AMANHI) prospective cohort study. J Glob Health. 2017;7(2):021201.

20. He K, Zhang X, Ren S, Sun J. Deep Residual Learning for Image Recognition2015 December 01, 2015:[arXiv:1512.03385 p.]. Available from: https://ui.adsabs.harvard.edu/abs/2015arXiv151203385H.

21. Zhou Z, Mahfuzur Rahman Siddiquee M, Tajbakhsh N, Liang J. UNet++: A Nested U-Net Architecture for Medical Image Segmentation2018 July 01, 2018:[arXiv:1807.10165 p.]. Available from: https://ui.adsabs.harvard.edu/abs/2018arXiv180710165Z.

