## supplementary material for "Development and external validation of an ultrasound image-based deep learning model to estimate gestational age in the second and third trimesters of pregnancy using data from Garbh-Ini cohort: a prospective cohort study in North Indian population"

Supplementary Table 1: Comparison of baseline characteristics of participants included in this study with all the participants of Garbh-Ini

| **Variables** | **Median (IQR) or N (%)** | **Median (IQR) or N (%)** |
|---|---|---|
| Age | 23.0 (21.0, 26.0) | 23.7(21.0, 26.6) |
| Body Mass Index (BMI)<br>· Underweight<br>· Normal<br>· Overweight<br>· Obese | <br>465 (21%)<br>1,429 (65%)<br>255 (12%)<br>45 (2.1%) | <br>2150 (27%)<br>4881 (61%)<br>819 (10%)<br>133 (1·7%) |
| Parity:<br>· Nulliparous<br>· Parous | <br>1,122 (51%)<br>1,072 (49%) | <br>4042 (50·6)<br>3958(49.4) |
| Appropriate for gestational age (AGA)<br>Small for gestational age (SGA) | 1,173 (62%)<br>665 (35%) | 3747(61.5%)<br>2345(38.4%) |
| Preterm Birth<br>Term Birth | 275 (13%)<br>1,820 (87%) | 753(12.3%)<br>5339(87.6%) |
| Occupation:<br>· Unemployed<br>· Employed | <br>2,014 (92%)<br>179 (8%) | <br>7336 (91·8%)<br>653(8.1%) |
| Education:<br>· Illiterate<br>· Literate | <br>406 (19%)<br>1,788 (81%) | <br>1602(20.1%)<br>6388(79.2%) |

Supplementary Table 2: Showing the performance (MAE) in days of the image regression models trained on the head, abdomen, and femur images across 18-20, 20-30 and 30+ weeks window.

| Anatomy | MAE | Test MAE 18-20 w | Test MAE 20-30 w | Test MAE 30+ w |
|---|---|---|---|---|
| Head | 3.51 [3.25 - 3.77] | 2.99 [2.75 – 3.22] | 5.06 [4.18 - 5.94] | 3.62 [3.09 – 4.20] |
| AC | 7.15 [6.60 – 7.70] | 4.06 [3.72-4.40] | 11.30 [8.79 - 13.81] | 7.47[6.75 – 8.20] |
| Femur | 6.38 [5.88 – 6.88] | 4.09 [3.72 – 4.45] | 9.56 [6.89 – 12.23] | 7.25 [6.49 – 8.02] |

Supplementary Table 3: Performance of GAUGE model in internal and external test sets before and after applying our conformal prediction (CP) algorithm.

| | Overall MAE | 18-20 wks MAE | 20-30 wks MAE | 30+ wks MAE |
|---|---|---|---|---|
| Internal test | 3.38 [3.06 – 3.70] | 2.45 [2.25 – 2.66] | 11.63 [7.86 – 15.41] | 3.14 [2.90 –3.40] |
| Internal test with CP | 2.84 [2.65 – 3.03] | 2.40 [2.16 – 2.62] | 5.62 [4.68 – 6.55] | 2.98 [2.72 – 3.23] |
| External test | 9.04 [7.99 –10.08] | 4.96 [3.70 – 6.22] | 10.04 [8.77 – 11.30] | 12.97 [9.52-16.41] |
| External test with CP | 5.96 [5.22 – 6.71] | 3.71 [3.04 – 4.38] | 7.80 [6.37 – 9.23] | 4.97 [2.22 – 7.70] |

Supplementary Table 4: Comparison of GAUGE, Hadlock and INTERGROWTH-21st models MAE (days) in SGA and AGA groups reported overall and in 18-20, and 30+ weeks windows.

| GA window | Group | GAUGE | Hadlock | INTERGROWTH-21st |
|---|---|---|---|---|
| Overall | AGA | 2.66 [2.45 - 2.87] | 4.41 [4.06 - 4.76] | 4.34 [3.99 - 4.69] |
| | SGA | 2.94 [2.66 - 3.22] | 6.51 [5.74 - 7.21] | 4.55 [4.20 - 5.04] |
| 18-20 | AGA | 2.38 [2.03 - 2.73] | 2.52 [2.17 - 2.87] | 2.59 [2.38 - 2.87] |
| | SGA | 2.45 [2.03 - 2.87] | 2.80 [2.38 - 3.15] | 2.73 [2.31 - 3.08] |
| 30+ | AGA | 2.87 [2.45 – 3.22] | 5.67 [5.18 – 6.16] | 5.53 [4.97 - 6.09] |
| | SGA | 3.01 [2.59 - 3.5] | 9.24 [8.26 -10.22] | 5.95 [5.32 - 6.58] |

Supplementary Table 5: Comparison between baseline head GA regression model and GAUGE model in 18-20, 20-30 and 30+ week window.

| Model | MAE | Test MAE 18-20 w | Test MAE 20-30 w | Test MAE 30+ w |
|---|---|---|---|---|
| Head Image Regression | 3.51 [3.25 -3.77] | 2.99 [2.75-3.22] | 5.06 [4.18 - 5.94] | 3.62 [3.09 – 4.20] |
| GAUGE | 2.84 [2.64 - 3.03] | 2.40 [2.16 – 2.62] | 5.62 [4.73 – 6.50] | 2.98 [2.70 – 3.25] |

Supplementary Fig. 1: Plot showing relationship between absolute difference in error (true – predicted GA) and transformed Mahalanobis distance in external and internal test datasets. Trend lines show a linear regression fit along with

95% confidence interval (shaded area). Data is binned (10 bins) and 95% CI for each bin are visualized using vertical lines.

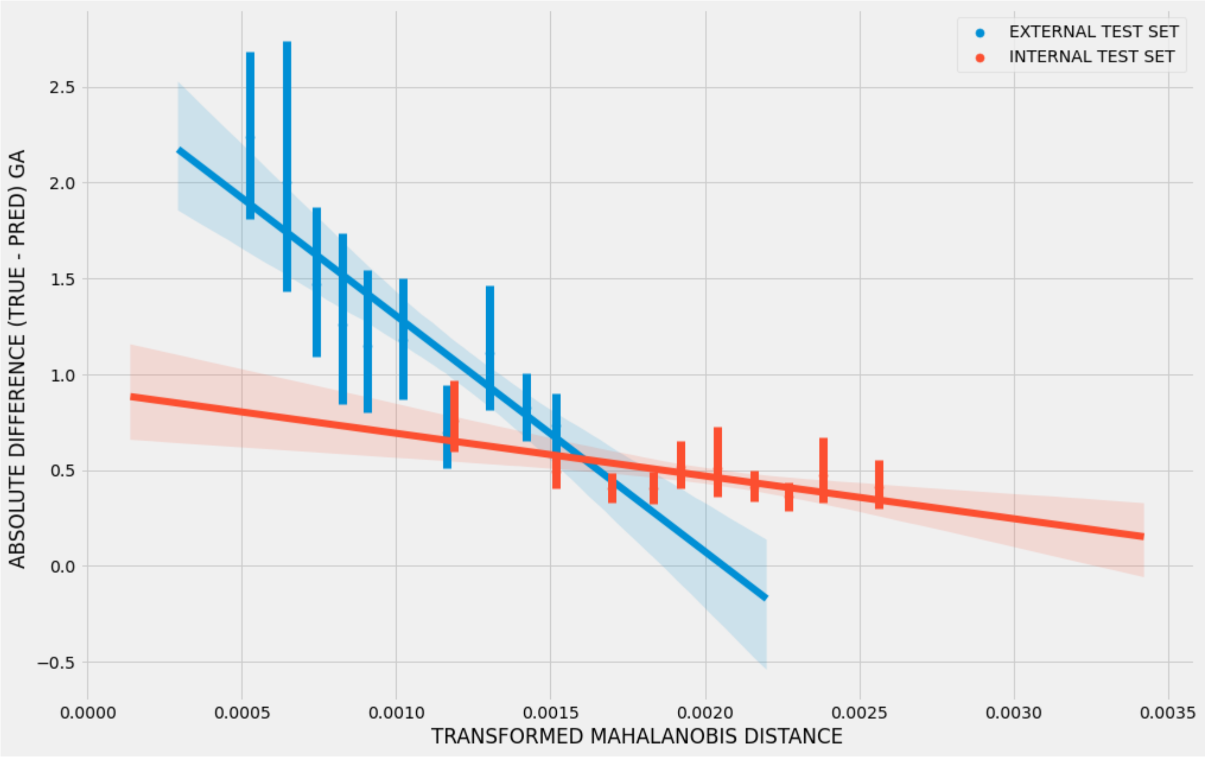

Supplementary Fig. 2: Showing the distributions of transformed Mahalanobis distance in the training, internal and external test datasets

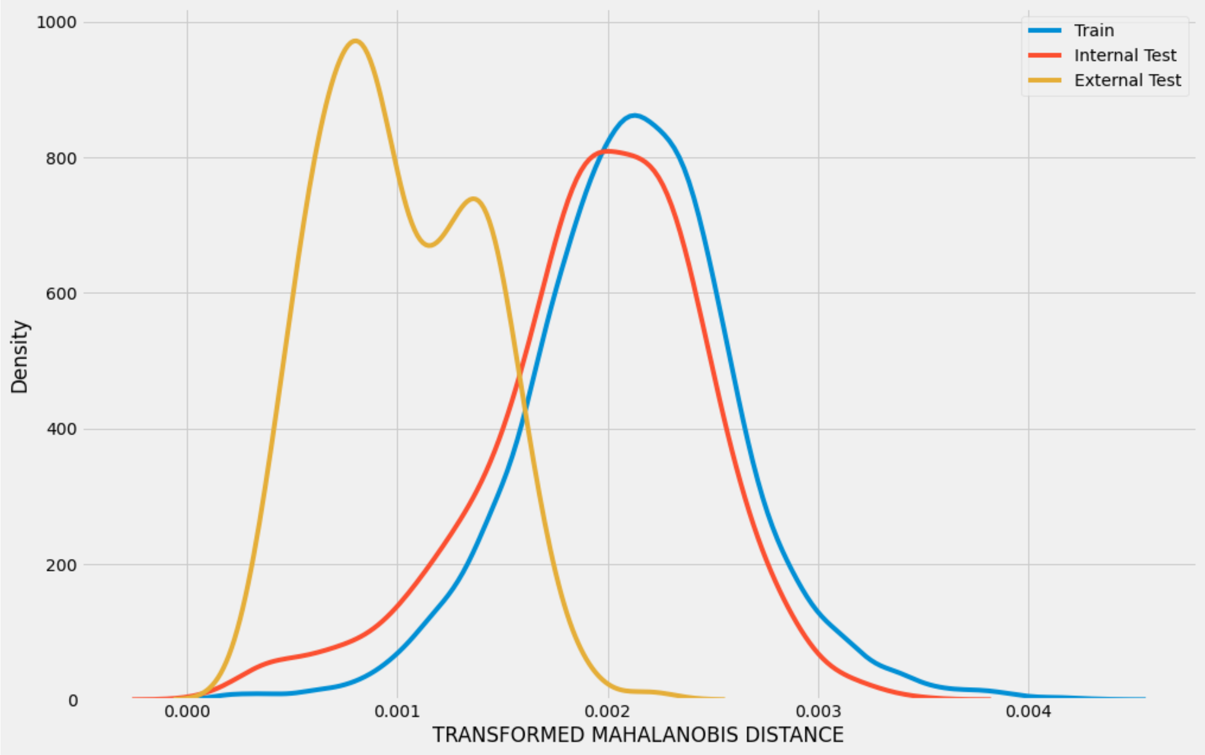


Supplementary Fig. 3: Regression Activation Maps (RAM) showing the model's dependency on finer details within the head region.

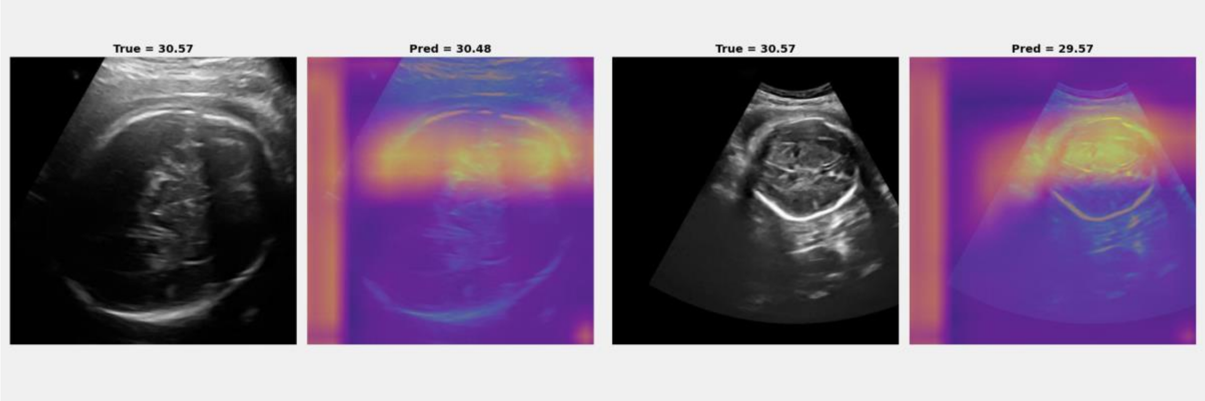

Supplementary Fig. 4: Boxplots showing the effect of different level of image resizing and blurring on the Mahalanobis distance.

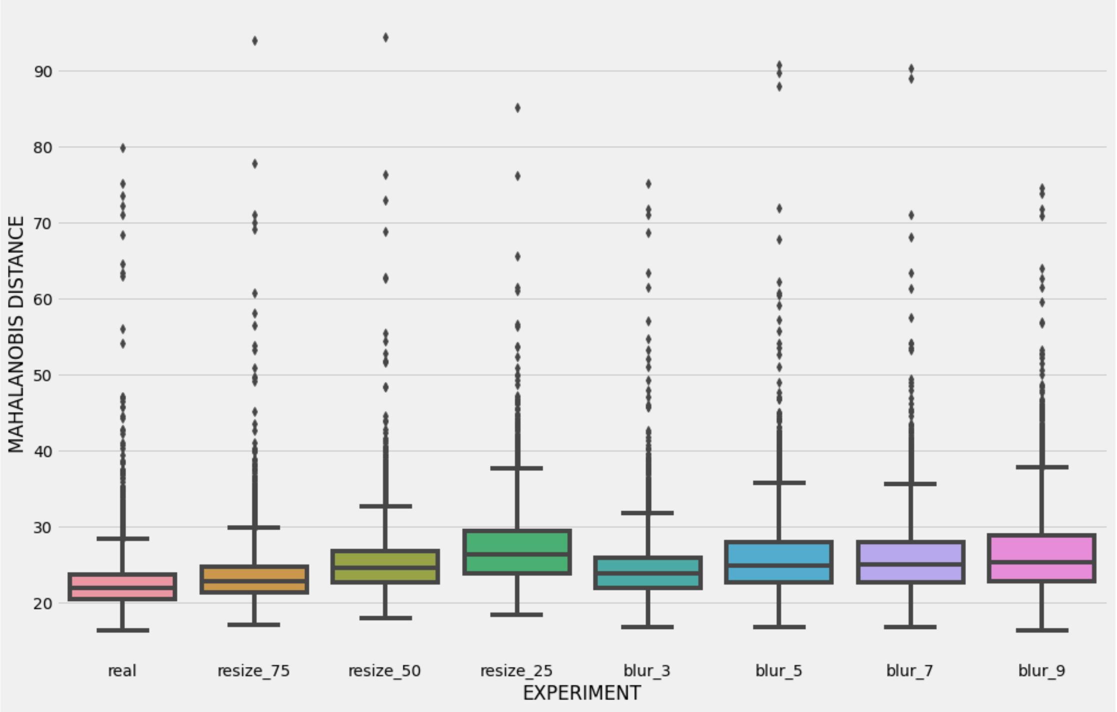

Supplementary Fig. 5: Plots showing that Mahalanobis distance attained normal distribution after ($y = 1/x^2$) transformation

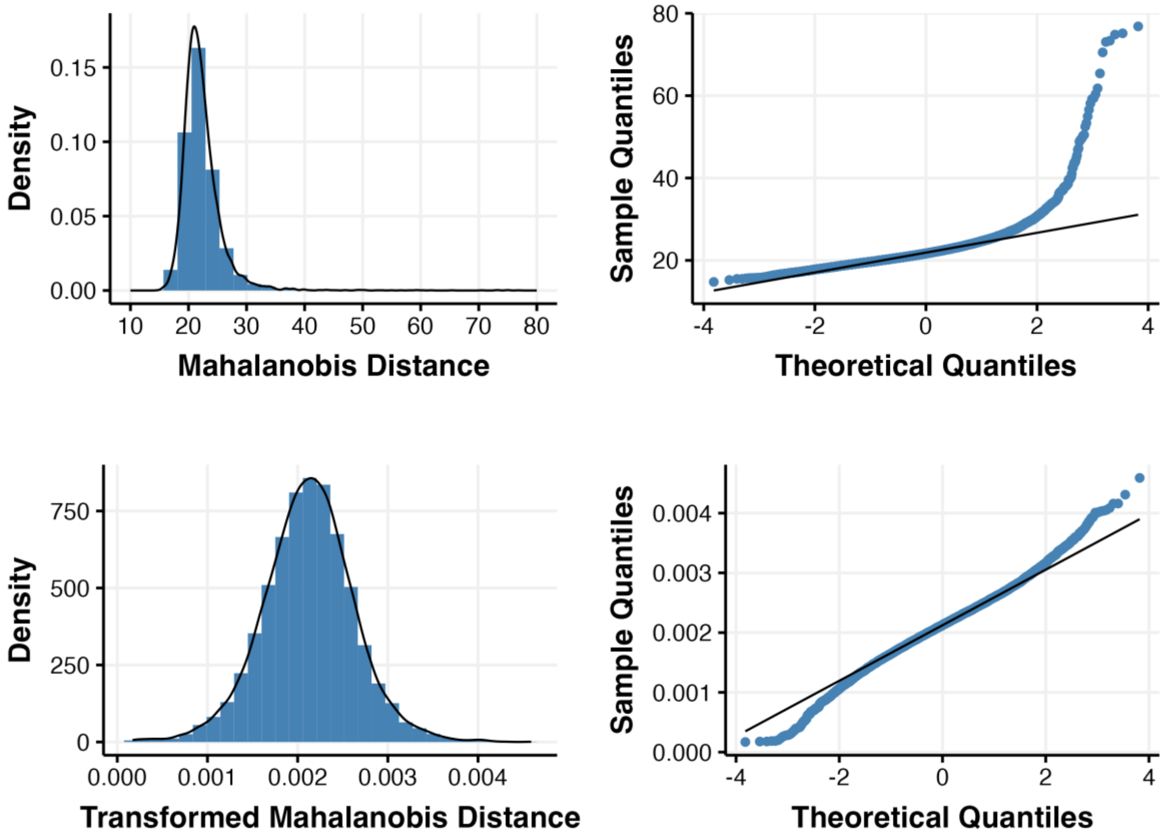


### OCR system for finding head, abdomen, and femur images:

To find images belonging to head, abdomen and femur class, an OCR based pipeline was designed. In the first stage, the image is cropped to remove machine buttons and other machine metadata. The cropped image is forwarded to thresholding stage where a mask containing only pixels of yellow colour is produced while other pixels are set to zero. Pixels of only yellow colour are taken because radiologists used the colour to annotate the images.
To increase the clarity of thresholded pixels, morphological operations are applied on the mask and is passed to the pytesseract OCR function to detect the text in the image.
To maximize detection,we designed anatomy-specific regular expressions that analyze the global pattern of the text and detect cases where some letters are missing or detected wrongly. The final text is used to sort the images and copy them in anatomy specific folders.
